# A large language model-assisted workflow for generating a living evidence base for climate-sensitive foodborne disease

**DOI:** 10.64898/2026.07.04.26357263

**Authors:** Richard Elson, K. Marie McIntyre, Megan B. Hardingham, Thomas Luechtefeld, Iain R. Lake

**Affiliations:** School of Environmental Sciences, University of East Anglia Norwich NR4 7TJ, UK; National Institute for Health and Care Research Health Protection Research Unit in Gastrointestinal infections, University of East Anglia and Newcastle University, UK; Modelling, Evidence and Policy group, School of Natural and Environmental Sciences, Newcastle University, UK; Insilica LLC, Bethesda, MD, United States

## Abstract

Climate change is altering environmental conditions that influence foodborne disease transmission, yet traditional systematic reviews cannot keep pace with expanding evidence. We assessed whether an LLM-assisted workflow could generate a rapid, repeatable, and policy-relevant living evidence base for climate-sensitive foodborne disease.

We combined structured PubMed searches (2010–2023), gold-standard human labelling, and iterative refinement of a GPT-4-Turbo-based auto-labeller within the SysRev platform. Pathogens of public-health importance in England were selected a priori. Model performance was evaluated against human reviewers using recall, precision, specificity, accuracy, and balanced accuracy.

The refined inclusion model achieved 89·2% recall, 59·2% precision, 84·5% specificity, and 85·4% accuracy across 1,044 screened abstracts, identifying 436 studies for inclusion. Post-hoc re-evaluation of discordant abstracts showed that records excluded by the model but included during initial human screening did not meet the refined inclusion criteria. Frequently identified climate exposures included rainfall, temperature, seasonality, and humidity; norovirus, *Salmonella*, *Campylobacter*, and *Cryptosporidium* were the most common pathogens.

An LLM-assisted workflow can generate living evidence for climate-sensitive foodborne disease with high recall and improved screening consistency. The approach is scalable, auditable, and suitable for secure institutional environments, supporting horizon scanning and climate-health risk assessment.

## Introduction

Foodborne pathogens are sensitive to climatic drivers such as temperature, precipitation, humidity, and seasonal variability, which affect pathogen survival, contamination pathways, and human exposure. Recent reviews report consistent positive associations between ambient temperature and gastrointestinal infections across multiple regions, with both linear and non-linear effects observed^1^. In Europe, nearly half of important human pathogens have foodborne or waterborne transmission routes, and foodborne pathogens show some of the strongest associations with climatic drivers^2^. As climate change advances, the burden of foodborne disease is expected to increase, disproportionately affecting socially and geographically marginalised populations^3^. Socio-economic disparities in gastrointestinal infection risk, particularly in young children^4–10^, underscore the importance of equity-aware decision-support systems^11^. Policymakers therefore need climate-informed surveillance systems^3^.

Public health agencies and other policy makers increasingly require rapid, contemporary, repeatable assessments of evidence on climate-sensitive health risks to guide adaptation strategies and strengthen resilience. Traditional systematic reviews enable rigorous synthesis but are too slow and resource-intensive to match the accelerating pace and heterogeneity of climate–health evidence. National-scale assessments can require screening tens of thousands of records and manually reviewing many thousands of abstracts. For example, the UK Health Security Agency’s 2023 climate-health assessment, required screening of more than 78,000 articles and manual review of over 6,500 abstracts to assess the likely impacts of climate change on infectious diseases in the UK^12^. Global health assessments similarly highlight the complex interaction between disease, socio-economic factors and climate mitigation policies and the need for more agile evidence systems^13^. This has catalysed interest in machine-learning-assisted approaches, including LLMs, to accelerate or automate key stages of the review pipeline while maintaining transparency and human oversight.

Although evaluations show promising performance for LLM-assisted screening, prioritisation, and risk-of-bias assessment, most studies are proof-of-concept and few have been tested within operational public-health workflows or designed to support living reviews. We address this gap by developing and evaluating an LLM-assisted workflow to generate a reproducible, low-cost, and easily updated evidence base for climate-sensitive foodborne disease.

This study aims to design, implement, and evaluate a machine-learning-assisted workflow that supplements and streamlines manual searching and screening to generate a living evidence base on climate-sensitive foodborne pathogens, with a focus on use within institutional decision-support settings.

## Methods

An overview of the process followed is presented in Figure 1. We conducted a methodological evaluation of an LLM-assisted screening workflow embedded in the SysRev platform, a web application designed to facilitate data curation and systematic evidence reviews^14^. The case study focused on foodborne pathogens of public-health importance in England selected for plausible climate sensitivity and policy relevance.

**Figure 1.**
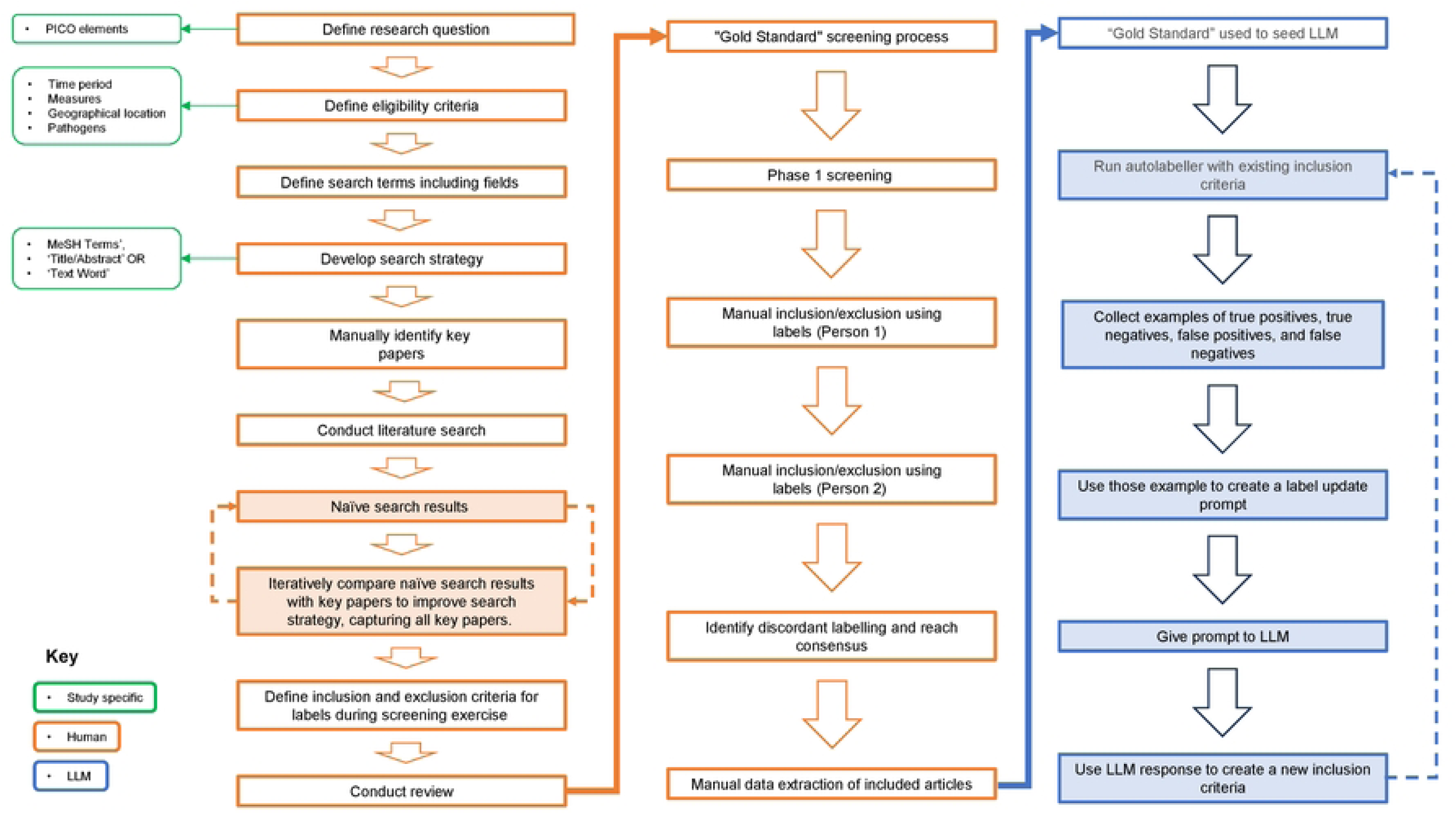

## Pathogen selection

Pathogens were selected from the 2023 UKHSA climate–health assessment^12^ and statutory notifications, including *Campylobacter* spp., *Cryptosporidium* spp., *Listeria monocytogenes, Salmonella* spp. and verocytotoxigenic *Escherichia coli* (VTEC)^15^. *Yersinia enterocolitica* was included as an emerging foodborne pathogen and norovirus was included due to its high disease burden and marked seasonality. Pathogens rare in the UK (including *Vibrio cholerae, Bacillus anthracis, Clostridium botulinum* and *Entamoeba histolytica*), predominantly non-foodborne (*Shigella spp., Giardia lamblia, Hepatitis A*), or typically associated with intoxication rather than infection (*Bacillus cereus, Clostridium perfringens*) were excluded to focus on plausible climate–food safety interactions.

## Information sources and search strategy

We searched PubMed (via SysRev) for literature published from Jan 1, 2010, to Dec 31, 2023, using English search terms (including translated foreign-language publications indexed by PubMed). Search terms combined MeSH and text words. For pathogens, climate/weather drivers (e.g., temperature, precipitation, humidity, extreme events, seasonality), and human health, joined as: [pathogen terms] AND [climate driver terms] AND [humans]. The full search string is provided in the supplementary material SM 1.

## Eligibility criteria

### Inclusion

- studies describing human disease caused by pathogens of interest and
- explicitly linking climate, weather, or seasonality to human disease outcomes.

### Exclusion

- animal-only studies; laboratory/mechanistic studies without epidemiological outcomes and
- studies without climate-related exposure.

Three reviewers independently screened an initial set of 365 abstracts against the eligibility criteria and extracted structured information (Supplementary Material SM 2). Disagreements were resolved by consensus to produce a gold-standard dataset used to seed and evaluate the auto-labeller. Inter-rater agreement statistics are available via the link in supplementary material SM 3.

SysRev’s auto-labeller (GPT-4-Turbo) was configured to classify abstracts against user-defined labels. Initial prompts were constructed directly from the inclusion criteria.

Drawing on emerging hybrid systematic review methodologies (e.g.^16,17^), we implemented an agent-driven refinement process to:

(1) run the auto-labeller on the gold-standard set;
(2) identify true/false positives and negatives;
(3) sample misclassified abstracts;
(4) feed these examples back to the LLM to update the inclusion criteria; and
(5) regenerate the inclusion prompt and re-run the auto-labeller.

Iterations continued until performance stabilised. Examples of prompt evolution are provided in the supplementary material and SM 4.

### Evaluation metrics

We evaluated recall, precision, specificity, accuracy, and balanced accuracy on abstracts screened by a single human reviewer or where multiple reviewers agreed on all extracted fields. Performance was compared narratively with recent evaluations of LLM-assisted screening. Final performance metrics were calculated on abstracts screened using the stabilised inclusion criteria, separate from those used during iterative prompt refinement.

## Results

### Performance of the inclusion model

Iterative refinement substantially improved model performance. Using the final inclusion criteria, recall was 89·2%, precision 59·2%, specificity 84·5%, and accuracy 85·4%, corresponding to 58 true positives, 218 true negatives, 40 false positives, and 7 false negatives across the evaluation set (n=1,044 abstracts) (Figure 2).

**Figure 2.**
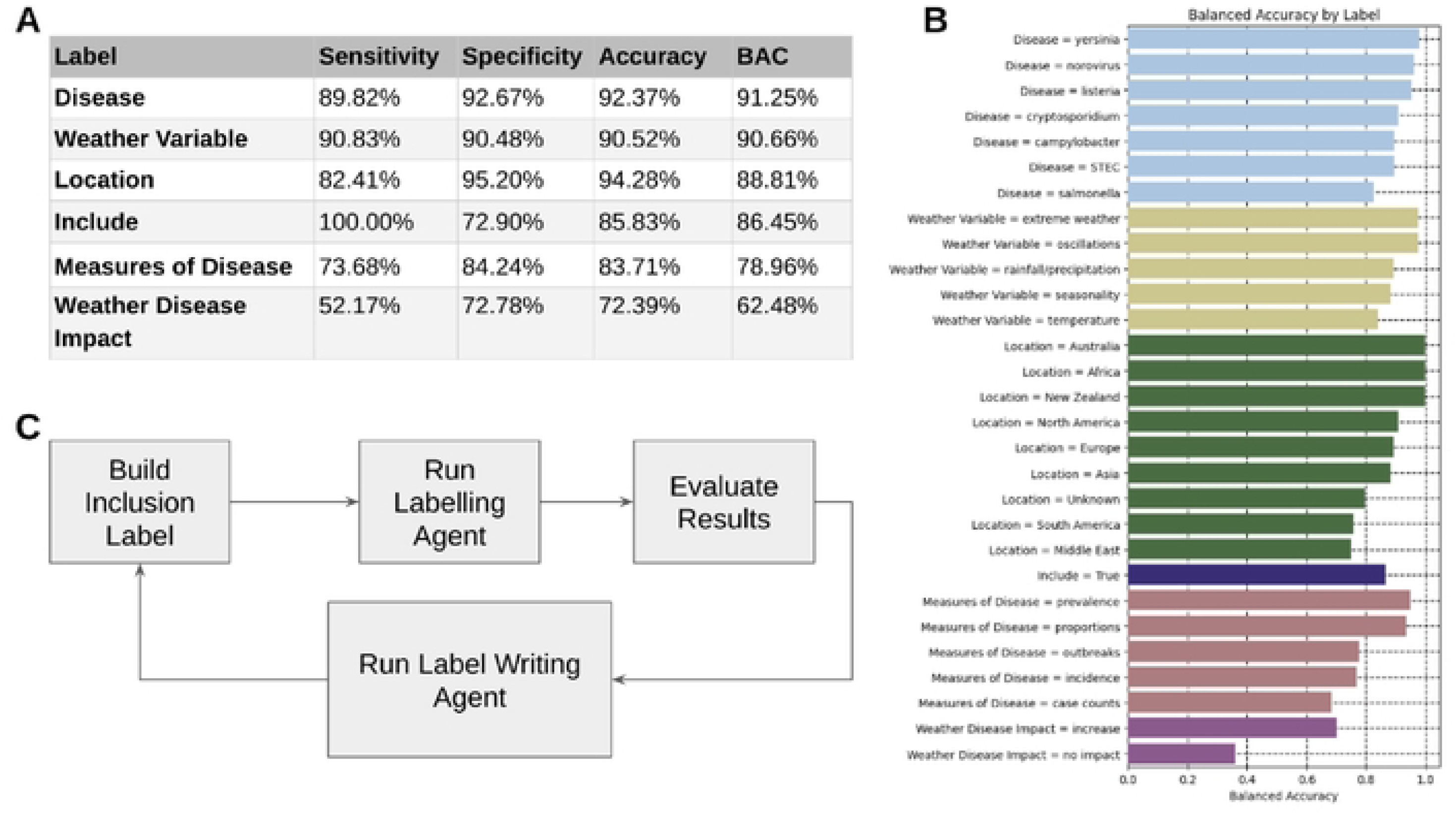
A. GPT4-Turbo p Figure 1A shows the performance metrics for each SysRev label evaluated in this study. For categorical variables with n1ore than two possible responses, mean perforn1ance values are presented. For the binary Include label, sensitivity, specificity, accuracy, and balanced accuracy, vere calculated. Figure 2B displays balanced accuracy for all extracted values, illustrating variation in model performance across different classification tasks. Figure 2C sumn1arises the iterative refinement process for the Include label. The workflow involved:

1. Running the auto-labeller,
2. Identifying misclassified abstracts,
3. Sampling false positives and false negatives,
4. Feeding these exan1ples back to the LLM, and
5. Updating the inclusion criteria before rerunning the model. This process was repeated until performance stabilised. Examples of the update script and prompt evolution are provided in Appendices 2 and 3.

During post-hoc adjudication, abstracts excluded by the auto-labeller but included during initial human screening were re-evaluated against the final, refined eligibility criteria and judged not to meet those criteria. This process highlights how iterative clarification of inclusion definitions can improve screening consistency in hybrid human–LLM workflows.

### Proportion and characteristics of included studies

The auto-labeller identified 436/1,044 (41·7%) abstracts as meeting the refined inclusion criteria. Figure 3 summarises the distribution of included articles by location, weather variable, and disease, with locations represented by fewer than 10 articles omitted from the analysis.

**Figure 3.**
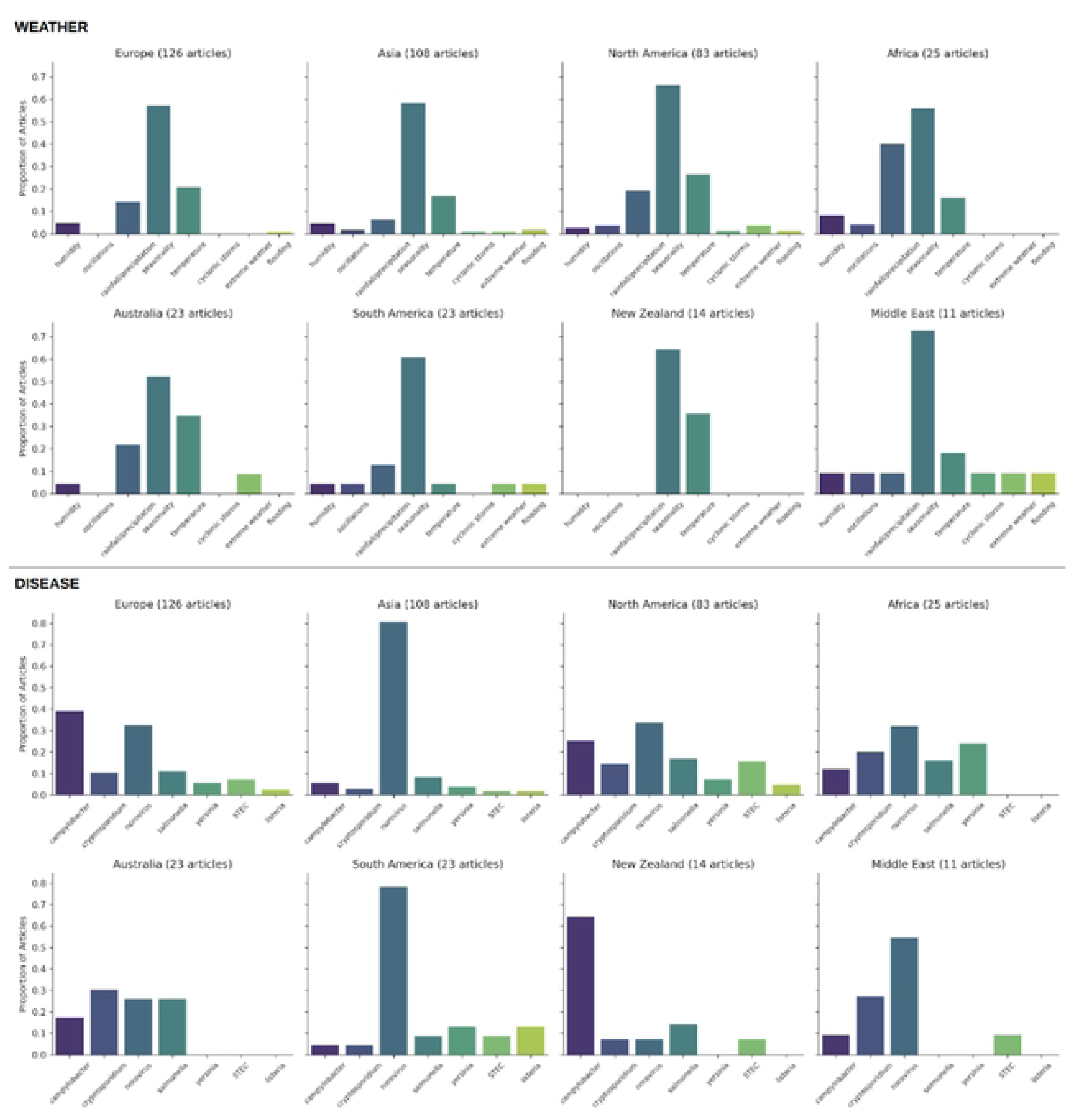
Characteristics of included studies identified by the LLM-assisted workflow. The upper panel shows the proportion of articles with each weather type by location as predicted by GPT4-Turbo. The lower panel shows the proportion of articles with each disease type by location as predicted by GPT4-Turbo.

Among the studies included, the most frequent climate variables were rainfall/precipitation, temperature, seasonality, and humidity (Figure 3). The most represented pathogens were norovirus, Salmonella spp., Campylobacter spp., and Cryptosporidium spp (Figure 3.). Geographically, included studies were most often from Europe, Asia, and North America (Figure 3).

## Discussion

In this study, we developed and evaluated a workflow that integrates structured literature searching, gold-standard human labelling, and iterative refinement of a large language model (LLM)–assisted auto-labeller to support systematic evidence synthesis. The final model achieved high recall (89.2%) and overall accuracy (85.4%), indicating strong performance in identifying relevant studies within a large and heterogeneous evidence base.

These findings demonstrate that LLM-assisted approaches can be effectively embedded within real-world screening workflows to reduce manual burden while maintaining sensitivity to relevant literature. Importantly, iterative refinement of the inclusion criteria using misclassified examples appeared to improve classification consistency, highlighting the value of hybrid human–LLM approaches rather than fully automated pipelines.

The workflow also addresses a recognised limitation of conventional systematic reviews, which are often resource-intensive and difficult to update in rapidly evolving fields such as climate–health. By supporting repeatable and auditable screening processes, LLM-assisted approaches offer a potential foundation for “living” evidence systems that can be updated as new literature emerges.

The high recall observed is particularly important in the context of systematic review screening, where missing relevant studies is typically considered more problematic than reviewing additional irrelevant records. The lower precision (59.2%) observed here therefore reflects a deliberate trade-off: prioritising sensitivity to minimise false negatives while retaining manageable levels of false positives for downstream human review.

In practice, this suggests that LLM-assisted screening could be used to triage large volumes of literature, substantially reducing reviewer workload while preserving the integrity of evidence identification. Rather than replacing human reviewers, the workflow is best viewed as augmenting decision-making by narrowing the candidate evidence set and standardising application of eligibility criteria.

Although formal time savings were not quantified, the workflow is expected to reduce the number of records requiring full manual screening by prioritising likely relevant studies.

## Comparison with other studies

Our findings are consistent with recent evaluations of LLM assisted screening^19–21^ that report strong recall and meaningful workload reductions for LLM-assisted screening and review automation^22^. A key contribution of this study is the explicit use of an iterative refinement process that updates inclusion criteria based on misclassified examples. This approach extends prior work by demonstrating how model performance can be improved within an operational workflow, rather than relying on static prompts or one-off evaluations^16,17^, and aligns with recommendations emphasising transparency, auditability, and human oversight^18^.

In addition, the application to climate-sensitive foodborne disease provides a realistic test case, characterised by heterogeneous exposures, varied study designs, and complex terminology. This strengthens the relevance of the findings for applied public health evidence synthesis.

The geographical distribution of included studies, dominated by Europe, North America, and Asia, reflects global research activity and the concentration of climate–health surveillance systems. This pattern highlights persistent geographic inequities in climate-health evidence generation, with implications for global resilience and adaptation planning.

The most frequently identified climate variables, rainfall, temperature, seasonality, and humidity, align with established climate drivers of foodborne disease, consistent with previous assessments of climate sensitivity^2^. Systematic evidence also shows that heavy rainfall and flooding consistently elevate diarrhoeal disease risk, particularly where water and sanitation infrastructure is vulnerable^23^.

The prominence of norovirus reflects its high global burden and strong seasonal patterns, while UK-focused reviews highlight strong temperature associations for Salmonella and complex, multi-pathway climate sensitivities for Campylobacter^3^. High-resolution analyses of Campylobacter in England and Wales further demonstrate non-linear temperature effects and region-specific patterns, reinforcing the need for pathogen-specific modelling^24^.

Climate–health relationships are inherently non-stationary, with changing exposure distributions, emerging hazards, and context-specific vulnerabilities. Static systematic reviews rapidly become outdated in this setting, limiting their utility for adaptation planning and horizon scanning. Living evidence approaches are therefore particularly important for climate-sensitive infectious diseases, where timely synthesis is needed to inform surveillance priorities, risk assessment, and policy responses under uncertainty. The workflow presented here is designed to support this adaptive evidence function rather than to replace conventional systematic reviews for causal inference or quantitative synthesis.

## Implications for decision-support and climate resilience

Public health agencies increasingly require rapid, repeated, and potentially policy-relevant assessments to support climate adaptation planning, needs that traditional systematic reviews cannot meet. The workflow presented here offers a low-cost, scalable, and reproducible alternative that can be updated frequently to support horizon scanning, early warning, evidence tracking, and decision-support applications for climate–health risk assessment.

These capabilities align with calls for dynamic, data-driven decision-support systems in climate-health^11^. They also parallel broader advances in AI for environmental and planetary health, where machine-learning models are increasingly used to generate high-resolution exposure surfaces and evaluate complex multi-pollutant health effects^25^. Integrating LLM-assisted evidence synthesis with such AI-enabled environmental data systems could support more holistic climate-health decision-support frameworks, enabling real-time updates as new evidence emerges.

## Strengths and limitations

This study has several strengths. First, it integrates human expertise with LLM-assisted classification and interoperability with PubMed in a transparent and reproducible workflow, rather than treating the model as a standalone solution. Second, performance was evaluated using multiple standard metrics, allowing a balanced assessment of sensitivity and specificity. Third, the workflow is designed to be adaptable, with inclusion criteria explicitly defined and iteratively refined. This supports consistency in screening decisions and facilitates reuse in other domains. Finally, the application to a large, nationally relevant topic demonstrates feasibility in a realistic public health context, where timely evidence synthesis is increasingly required.

Several limitations should be considered when interpreting these findings.

First, screening was conducted using abstracts rather than full texts. While this reflects common practice in early-stage systematic review screening, it may limit generalisability to later stages of evidence synthesis where full-text assessment is required.

Second, the workflow was implemented using a single LLM model and platform. Although the overall approach is designed to be model-agnostic, performance may vary across different LLMs, prompting strategies, or deployment environments.

Third, model performance was evaluated within a single domain-specific dataset. While the inclusion criteria were refined iteratively, external validation in other topic areas would strengthen confidence in generalisability.

Fourth, the observed precision indicates that a substantial proportion of records identified as relevant would require exclusion at later stages. Although this is acceptable in screening contexts prioritising recall, it reinforces the continued need for human oversight.

Finally, this study focuses on the identification of relevant studies and does not address downstream stages of evidence synthesis, such as data extraction, risk-of-bias assessment, or meta-analysis.

As a methodological evaluation, this study does not aim to generate definitive causal conclusions but to assess the feasibility of maintaining timely, policy-relevant evidence under conditions of rapid evidence growth. Substantial mechanistic uncertainty for several foodborne pathogens underscores the need for deeper extraction and synthesis in future automated pipelines^3^. Future work should explore automated extraction of epidemiological measures, bias assessment, meta-analysis, integration with climate-risk modelling frameworks and deployment within secure, firewalled environments.

## Implications for policy decision-support

Public-health agencies need rapid, repeatable, and policy-ready assessments to inform climate-adaptation planning and horizon scanning. This workflow provides a low-cost and scalable foundation for living reviews and can be deployed in firewalled environments used by government agencies. Integrating LLM-assisted evidence synthesis with environmental data systems and climate-risk models could enable more holistic, real-time decision-support frameworks.

Future work should evaluate the performance of similar workflows across a wider range of topics and incorporate external validation datasets to assess robustness. Extending the approach to full-text screening and downstream stages of systematic review pipelines would also be valuable.

In addition, quantifying reductions in screening time and resource requirements would provide important evidence for adoption in operational settings. Integration with existing systematic review platforms and secure institutional environments may facilitate translation into routine use.

Finally, the combination of LLM-assisted screening with emerging data extraction and synthesis tools offers a pathway towards more adaptive and continuously updated evidence systems, particularly in areas such as climate–health where the evidence base is rapidly evolving.

## Conclusion

We demonstrate that an LLM-assisted workflow combining structured searches, human expertise, and iterative refinement can achieve high recall and robust overall performance in identifying climate-sensitive health evidence. While not a substitute for human judgement, this approach provides a low cost, scalable and reproducible method to support systematic evidence synthesis and may contribute to more efficient, updatable evidence systems in public health.

## Contributors

All authors contributed to study design. RE, IRL, KMM and MH developed the search strategy and performed initial screening. TL implemented the SysRev workflow and iterative prompt refinement. TL and RE conducted the analysis. RE, TL, KMM and IRL drafted the manuscript. All authors reviewed and approved the final version.

## Data sharing

Inclusion criteria, prompt evolution, and code for label updates are available in the Supplementary Material. De-identified screening data can be made available upon request for research purposes.

## Data Availability

Inclusion criteria, prompt evolution, and code for label updates are available in the Supplementary Material. De?identified screening data can be made available upon request for research purposes.

## Acknowledgements

We thank colleagues at the UK Health Security Agency and the UK Food Standards Agency for domain guidance and discussions on operational deployment. RE, MH, IRL and KMM are supported by the National Institute for Health and Care Research Health Protection Research Unit in Gastrointestinal infections, University of East Anglia and Newcastle University, UK.

## Declaration of generative AI and AI-assisted technologies in the manuscript preparation process

During the preparation of this work the author(s) used Microsoft 365 Co-Pilot in order to review this manuscript for journal style and content. After using this tool/service, the author(s) reviewed and edited the content as needed and take(s) full responsibility for the content of the published article.

## References

1. Ghazani M, FitzGerald G, Hu W, Toloo GS, Xu Z. Temperature Variability and Gastrointestinal Infections: A Review of Impacts and Future Perspectives. Int J Environ Res Public Health 2018; 15(4).

2. McIntyre KM, Setzkorn C, Hepworth PJ, Morand S, Morse AP, Baylis M. Systematic Assessment of the Climate Sensitivity of Important Human and Domestic Animals Pathogens in Europe. Sci Rep 2017; 7(1): 7134.

3. Lake IR. Food-borne disease and climate change in the United Kingdom. Environ Health 2017; 16(Suppl 1): 117.

4. Adams N, Byrne L, Rose T, et al. Sociodemographic and clinical risk factors for paediatric typical haemolytic uraemic syndrome: retrospective cohort study. BMJ Paediatr Open 2019; 3(1): e000465.

5. Adams NL, Byrne L, Rose TC, et al. Influence of socio-economic status on Shiga toxin-producing Escherichia coli (STEC) infection incidence, risk factors and clinical features. Epidemiol Infect 2019; 147: e215.

6. Adams NL, Rose TC, Hawker J, et al. Relationship between socioeconomic status and gastrointestinal infections in developed countries: A systematic review and meta-analysis. PLoS One 2018; 13(1): e0191633.

7. Adams NL, Rose TC, Hawker J, et al. Socioeconomic status and infectious intestinal disease in the community: a longitudinal study (IID2 study). Eur J Public Health 2018; 28(1): 134–8.

8. Rose TC, Adams N, Taylor-Robinson DC, et al. Relationship between socioeconomic status and gastrointestinal infections in developed countries: a systematic review protocol. Syst Rev 2016; 5: 13.

9. Rose TC, Adams NL, Barr B, et al. Socioeconomic status is associated with symptom severity and sickness absence in people with infectious intestinal disease in the UK. BMC Infect Dis 2017; 17(1): 447.

10. Rose TC, Adams NL, Whitehead M, et al. Neighbourhood unemployment and other socio-demographic predictors of emergency hospitalisation for infectious intestinal disease in England: A longitudinal ecological study. J Infect 2020; 81(5): 736–42.

11. Rocklöv J, Semenza JC, Dasgupta S, et al. Decision-support tools to build climate resilience against emerging infectious diseases in Europe and beyond. Lancet Reg Health Eur 2023; 32: 100701.

12. UK Health Security Agency. Climate change: health effects in the UK, 2023. Available at: https://www.gov.uk/guidance/health-effects-of-climate-change-hecc-report. Last accessed: 27th March 2026.

13. Patz JA, Frumkin H, Holloway T, Vimont DJ, Haines A. Climate change: challenges and opportunities for global health. Jama 2014; 312(15): 1565–80.

14. Bozada T, Jr., Borden J, Workman J, Del Cid M, Malinowski J, Luechtefeld T. Sysrev: A FAIR Platform for Data Curation and Systematic Evidence Review. Front Artif Intell 2021; 4: 685298.

15. Anonymous. The Health Protection (Notification) Regulations 2010 (SI 2010/659). 2010. Available at: https://www.legislation.gov.uk/uksi/2010/659/contents. Last accessed: 27th March 2026.

16. Wang S, Scells H, Zhuang S, Potthast M, Koopman B, Zuccon G. Zero-Shot Generative Large Language Models for Systematic Review Screening Automation. Lecture Notes in Computer Science (including subseries Lecture Notes in Artificial Intelligence and Lecture Notes in Bioinformatics); 2024. p. 403–20.

17. Ye A, Maiti A, Schmidt M, Pedersen SJ. A Hybrid Semi-Automated Workflow for Systematic and Literature Review Processes with Large Language Model Analysis. Future Internet 2024; 16(5).

18. UK Department for Science, Innovation and Technology. Capabilities and risks from frontier AI. A discussion paper on the need for further research into AI risk., 2023. Available at: https://www.gov.uk/government/publications/frontier-ai-capabilities-and-risks-discussion-paper/frontier-ai-capabilities-and-risks-discussion-paper. Last accessed: 27th March 2026.

19. Issaiy M, Ghanaati H, Kolahi S, et al. Methodological insights into ChatGPT’s screening performance in systematic reviews. BMC Medical Research Methodology 2024; 24(1).

20. Khraisha Q, Put S, Kappenberg J, Warraitch A, Hadfield K. Can large language models replace humans in systematic reviews? Evaluating GPT-4’s efficacy in screening and extracting data from peer-reviewed and grey literature in multiple languages. Research Synthesis Methods 2024.

21. Qureshi R, Shaughnessy D, Gill KAR, Robinson KA, Li T, Agai E. Are ChatGPT and large language models “the answer” to bringing us closer to systematic review automation? Systematic Reviews 2023; 12(1).

22. Scherbakov D, Hubig N, Jansari V, Bakumenko A, Lenert LA. The emergence of large language models as tools in literature reviews: a large language model-assisted systematic review. J Am Med Inform Assoc 2025; 32(6): 1071–86.

23. Levy K, Woster AP, Goldstein RS, Carlton EJ. Untangling the Impacts of Climate Change on Waterborne Diseases: a Systematic Review of Relationships between Diarrheal Diseases and Temperature, Rainfall, Flooding, and Drought. Environ Sci Technol 2016; 50(10): 4905–22.

24. Djennad A, Lo Iacono G, Sarran C, et al. Seasonality and the effects of weather on Campylobacter infections. BMC Infectious Diseases 2019; 19(1): 255.

25. Han D, Xu Y, Lin L, Meng X, Chen R, Kan H. Artificial Intelligence in Environment and Human Health: Progress, Opportunities and Challenges. Curr Environ Health Rep 2025; 12(1): 45.

